# Cannabis Use and Sleep Quality in Daily Life: a Daily Diary Study of Adults Starting Cannabis for Health Concerns

**DOI:** 10.1101/2022.01.19.22269565

**Authors:** Brenden Tervo-Clemmens, William Schmitt, Grace Wheeler, Megan Cooke, Randi M. Schuster, Sarah Hickey, Gladys Pachas, A. Eden Evins, Jodi Gilman

## Abstract

**Background:** Legalization of cannabis for medical uses has proceeded without well-controlled studies. Real world patterns of medical cannabis use are highly variable and rarely overseen by a physician. Smartphone assessments that capture ecologically valid patterns of medical cannabis use and health symptoms may help clarify risks and benefits.

**Methods:** As part of a larger, randomized trial (NCT03224468), adults (N=181) seeking cannabis for insomnia, pain, or anxiety or depressive symptoms were randomized to obtain a medical cannabis card immediately (MCC) or to a waitlist control (WLC) and completed 12-weeks of daily web-based surveys on cannabis use and three health outcomes: sleep, pain, and depressive symptoms.

**Results:** Completion rates in this long-term, daily survey design were high (median completed assessments: 72 out of 90 days). Daily reports of cannabis use were consistent with monthly interview assessments and urinalysis. The MCC group increased cannabis use frequency following randomization, while WLC did not. Within the MCC group, self-reported sleep quality was significantly higher on cannabis use days, compared to nonuse days. The MCC group displayed long-term sleep improvements, paralleled by increased cannabis frequency. Daily associations between cannabis use and self-reported pain or depressive symptoms were not significant.

**Conclusion:** Cannabis use is associated with same day improvements in self-reported sleep quality, but not pain or depressive symptoms, although sleep improvements occurred within the context of potentially risky increases in use. Long-term, web-based assessments of cannabis appear valid and feasible, providing a robust method for future real-world effectiveness studies with expanded and objective measures.

**Funding/Support:** This work was funded by R01DA042043; PI: JMG.

## Introduction

Access to cannabis is rapidly increasing in the United States as a growing number of states legalize medical and recreational cannabis (Goodman *et al*., 2020). Though patients increasingly seek clinician input on the use of medical cannabis to address chronic mental and physical health challenges, including with sleep, mood, and pain (Sarris *et al*., 2020)(Lintzeris *et al*., 2018), the efficacy of medical cannabis is inconclusive (Abrams, 2018)(Sarris *et al*., 2020). Recent systematic reviews suggest that relatively stronger evidence exists for medical cannabis improving sleep quality, relative to improvements in mood symptoms (e.g., anxiety and depression) or reducing chronic pain (Abrams, 2018)(Sarris *et al*., 2020)(Analgesia, 2021). Nevertheless, mixed evidence exists within each symptom domain and few studies have evaluated the effects of cannabis on multiple domains in the same study.

In the United States, the complex legal status of cannabis (federally prohibited but with individual state-level laws (Boehnke *et al*., 2019) (Mead, 2017)(Goodman *et al*., 2020) (Pacula *et al*., 2014)) and lack of federal oversight (i.e., Food and Drug Administration: FDA) and evaluation of dosage, safety, and efficacy (fda.gov) pose a challenge in determining potential therapeutic effects of cannabis. In routine practice, many, if not most, medical cannabis patients use cannabis without ongoing physician supervision (Sexton *et al*., 2016), lack clear guidelines for dosing (see (MacCallum and Russo, 2018)), and have the flexibility to purchase a wide range of cannabis products (Hazekamp *et al*., 2013) (Cranford *et al*., 2016) of potentially unclear or mislabeled chemical composition (Vandrey *et al*., 2015)(Gilman *et al*., 2021b). Real-world patterns of medical cannabis use are thus highly variable, patient specific, and may be influenced by familiarity and/or expectations for use that facilitate navigation of its complex legal status.

Pragmatic clinical trials, that for example randomize individuals to receive access to medical cannabis or to a waitlist control (Gilman et al., invited to resubmit, (Gilman *et al*., 2021a)) provide an opportunity to characterize the potential therapeutic effectiveness of cannabis under real-world conditions. Experience sampling studies (Csikszentmihalyi and Larson, 2014)(Kahneman *et al*., 2004) that capture patient-specific, in-the moment (ecological momentary assessment) or on the same day (daily diary) patterns of cannabis use (Verdoux *et al*., 2003) and physical and mental health symptoms through intensive longitudinal tracking via web-based devices (e.g., smartphones) can provide essential complementary data for these pragmatic medical cannabis studies. Owing to the complex legal status of medical cannabis and the relatively novel methodological requirements of intensive longitudinal assessment, however, limited work has used experience sampling designs (e.g., daily diary, ecological momentary assessment) in studies of medical cannabis. This is particularly true across the relatively longer time scales used in pragmatic, effectiveness studies amenable to medical cannabis (e.g., 12 weeks) compared to the shorter timescales most frequently used in experience sampling and naturalistic studies of cannabis (<1 month)(Goodhines *et al*., 2019)(Verdoux *et al*., 2003)(Buckner *et al*., 2012))(Schuster *et al*., 2016)(Budney *et al*., 2001). Further, few studies have validated intensive web-based cannabis self-report from experience sampling studies against field-standard interview-based assessments or urinalysis. As such, despite the alignment of experience sampling design to studies of medical cannabis, the feasibility and validity of this method remains unknown.

As part of a larger pragmatic clinical trial that randomized patients interested in medical cannabis to a medical cannabis card or waitlist control (Gilman et al., invited to resubmit, (Gilman *et al*., 2021a)), the current project reports exploratory analyses from a novel intensive longitudinal, daily diary design. Participants provided daily web-based, self-reports of medical cannabis use and symptoms of sleep, mood, and pain for the first 90 days of receiving access to a medical cannabis card or placement on the waitlist. To our knowledge, this is the first intensive longitudinal, experience sampling study (ecological momentary assessment or daily diary) of new medical cannabis use and the first long-term (>1 month) experience sampling study of cannabis use. The primary aims for this study were to 1) determine the feasibility and validity of intensive longitudinal assessment of cannabis use in adults starting cannabis use for health concerns and 2) examine the association between cannabis use and reported symptoms of sleep, mood, and pain across the multiple timescales afforded by the long-term, daily diary design (e.g., short-term same day effects, long-term change over several months).

## Method

### Participants

Participants were part of a single site clinical trial (NCT03224468; Gilman et al., invited to resubmit, (Gilman *et al*., 2021a)) that randomized participants to either obtain a medical cannabis card (MCC) in the community at the time of randomization or to wait 12 weeks before obtaining a medical cannabis card (waitlist control (WLC)). Participants were adults (18-65-years-old) with self-reported symptoms of insomnia, pain, or anxiety and/or depression (“mood”), were generally healthy, with no known, unstable major medical condition, who expressed an interest in obtaining a medical cannabis card but did not yet possess one, reported less than daily current cannabis use, and did not meet criteria for a cannabis use disorder.

Our initial sample consisted of the 186 participants randomized to MCC or WLC who participated in monthly assessments (Gilman et al., invited to resubmit, (Gilman *et al*., 2021a)). The final analytic sample (see below) for this study consisted of 181 of these participants. Participant demographics and baseline characteristics are presented in Table 1. See Gilman et al., invited to resubmit, (Gilman *et al*., 2021a) for study design, participant characteristics, and randomization. Study procedures were approved by Partners Human Research Committee. Participants provided informed consent and were financially compensated for participation.

**Table 1.** Stratification and Demographic and Baseline Assessments from Analysis Sample.

|  | MCC | WLC |
| --- | --- | --- |
| *Demographics* |  |  |
| N Participants | 102 | 79 |
| Presenting Problem |  |  |
| (N Affective \| Sleep \| Pain) | 44 \| 22 \| 36 | 37 \| 19 \| 23 |
| N Female \| Male \| Non-Binary | 68 \| 33 \| 1 | 50 \| 29 \| 0 |
| Age in years | 38.26 (14.31) | 36.69 (14.56) |
| Education years | 16.71 (2.31) | 16.27 (2.74) |
| *Baseline Assessments* |  |  |
| Cannabis Uses Per-Day | 0.50 (0.63) | 0.58 (0.61) |
| Athens Insomnia Scale Total | 9.71 (4.95) | 9.63 (4.40) |
| HADS-Anxiety Total | 7.50 (4.35) | 7.91 (4.34) |
| HADS-Depression Total | 5.01 (3.51) | 5.04 (4.05) |
| Brief Pain Inventory Worst Pain | 5.12 (2.30) | 5.52 (2.20) |

*Note.* No significant differences ( $p$ 's > .243) were found between groups on demographic measures or baseline assessments. Number in parentheses following quantitative variables in standard deviation. HADS, Hospital Anxiety and Depression Scale. See document for measure citations.

### Procedures

Following a screening visit to assess basic study eligibility, participants were randomized, stratified by sex, age, and presenting problem for which they were seeking medical cannabis (self-reported problems with sleep, pain or mood [anxiety or depression]) to either the MCC group, in which they were to obtain a medical cannabis card without delay, or to the WLC group, in which they agreed to wait 12-weeks before obtaining a medical cannabis card. Study staff did not provide medical cannabis cards or cannabis products. Rather, participants randomized to the MCC group were instructed that they could obtain a medical cannabis card in the community without delay to participate in the study, and in detailed subsequent analyses, we determined changes in their cannabis usage via biochemically verified assessments. Owing to the expected dropout from the MCC group due to financial and logistic challenges inherent in obtaining a medical cannabis card, participants were randomized 2:1 MCC: WLC. Following randomization, participants completed an experimenter administered baseline visit where primary study variables, including cannabis use, sleep, anxiety and depression, and pain were assessed, and they received instructions on subsequent web-based assessments and daily self-reports. For the MCC group, the baseline visit was scheduled to be as close as possible to the receipt date of the medical cannabis card; if participants received the card prior to the baseline visit, they were instructed to not begin using their medical cannabis card until after the baseline visit (on average, MCC participants received their cards four days before the baseline visit; see Results for more discussion). Following the randomization and baseline assessment, participants were prospectively followed for the 12-week, randomized study period using 1) in-person or virtual experimenter administered visits (all experimenter administered visits became virtual in March 2020 due to COVID-19 pandemic) at 2-, 4-, and 12-weeks following randomization and 2) using daily web-based assessments of self-reported cannabis use, sleep, pain, depression. The current project reports the novel results of the daily web-based assessments.

### Measures

#### Baseline Substance Use, Psychiatric, Medical Symptoms

Symptoms of cannabis use disorder (CUD), a study exclusion criterion, were assessed by the CUD Checklist for DSM-5 (American Psychiatric Association, 2013). Baseline and monthly cannabis use frequency was assessed at each experimenter administered study visit via an interview with research staff where participants reported their cannabis use according to the following options: “Once or more per day”, “5-6 days a week”, “3-4 days a week”, “1-2 days a week”, “Less than once a week”, “Less than once every two weeks”. Baseline sleep, anxiety, depression, and pain symptoms were assessed via the Athens Insomnia Scale (Soldatos *et al*., 2000), Hospital Anxiety and Depression Scale (Snaith, 2003), and Brief Pain Inventory (Tan *et al*., 2004), respectively.

#### Daily Self-Reports

Participants provided daily reports on cannabis use and sleep, pain and depression symptoms via a secure web-based application, designed for this study, that was available via smartphone and computer. Participants used their own devices to complete the surveys. Participants were instructed to complete assessments at the same time every day; questions were based on the previous 24 hours. For cannabis use, participants first reported whether they had used cannabis (“Did you use Medical Marijuana Today”: “Yes” versus” No”) and if they had used cannabis that day, to report an approximation of the number of cannabis use occasions (“Please indicate the time(s) you used Medical Marijuana”; participants selected each hour of the day that they used cannabis [e.g., “6:00pm”,”7:00pm”, “8:00pm”, …], although hours were used as a memory aid and to provide a standard metric of occasion, but specific times were not saved.) Within this application, participants also provided daily reports, from 1 (low) to 10 (high) using sliders, on pain (“How much pain did you feel today on average”: “(1) No pain”, “(10) Extreme pain”), sleep (“How was your sleep quality last night”: “(1)Very poor”, “(10) Very good”). and depression (“How depressed did you feel today: “(1) Not at all”, “(10) Extremely”).

#### Urinalysis

At experimenter administered visits, whether in person or virtual, urine was collected for assessment of cannabinoids and their metabolites. Urine was collected in person for clinic-based visits and by mail for virtual visits. Concentration of THC, CBD, their primary metabolites, and 15 other cannabinoids in urine was assessed via high performance liquid chromatography with tandem mass spectrometry (see Supplemental S1 and (Gilman *et al*., 2021b) for more discussion).

#### Adverse Events

Adverse events are documented in the primary report from this trial (Gilman et al., invited to resubmit, (Gilman *et al*., 2021a)). No participants were withdrawn from the study due to an adverse event.

### Analysis

#### Feasibility and Validation of Daily Cannabis Reports

Feasibility analyses used descriptive statistics and per-participant and group-level data visualization to characterize the number of completed daily surveys. Validity analyses utilized linear regression to compare cannabis use reported in the daily diary design to cannabis use reported in field-standard, interview-based assessments, and cannabinoid metabolites from urinalysis.

#### Daily Diary Associations between Cannabis use and Physical and Mental Health Symptoms

Linear mixed effects models were used to examine associations between cannabis use and sleep, pain, and anxiety and depression symptoms across the 12-week daily diary period. Cannabis use and the effect of cannabis use on symptoms was examined at the within-person, day-level by comparing days with versus without reported cannabis use. To ensure our results reflected these within-person, same day effects, we covaried for between-person differences in the total number of cannabis use days. All models also included fixed and random effects (see below) of days since baseline to 1) account for person-specific changes in symptoms following randomization to MCC, when examining day-level effects and 2) to examine long-term changes in symptom expression following randomization. Random effects within mixed models, estimated for each participant, were an intercept, and slope effects for days since the baseline visit, the difference between use and nonuse days, and the interaction between days since randomization and use versus nonuse days. Per-participant autocorrelated (order 1) error structures, with respect to days since baseline, were also modeled. As a central focus of the project was on potential symptom relief from medical cannabis, analyses focused on same-day (i.e., zero lag) associations between cannabis use and health symptoms. For sleep quality, this meant the daily sleep measure (“How was your sleep quality last night”) was shifted forward one day (“lead”) with respect to the cannabis use metric, to model the effect of cannabis use on same-day (in the case of sleep, same-night) symptoms for all three outcomes.

Models were run with the three symptoms separately (sleep, pain, or depression) as dependent variables and cannabis use measures (use versus nonuse day, number of use occasions) as the independent variable. This model included age, years of education (see results for justification), and presenting problem (stratification variable: self-reported challenges with sleep, pain or mood) as covariates. In addition to the main effects, secondary analyses examined the associations between same day cannabis use and health symptoms in subgroups according to participant’s self-reported presenting problem that was used in stratification for randomization (see Procedure). Statistical inference was based on both effect size (e.g., differences in standard deviation units) and significance values (Cumming, 2014).

#### Probability of Cannabis Use Following Randomization

In order to better understand any potential longer-term changes in health symptoms following randomization, cannabis use frequency (use day versus nonuse day) was modeled as a dependent variable as a function of time since baseline, via generalized linear mixed effects models with a logit link function. Fixed effects again included participant age, years of education, and presenting problem as covariates. Random intercepts and a slope for days since baseline were included for each participant.

#### Sensitivity Analysis

Sensitivity analyses was performed restricting the analyses to only those MCC participants who had cannabinoid metabolites detected via urinalysis during at least one timepoint of the study period. Unless otherwise stated, the magnitude (effect size) and pattern of significance of primary results were unchanged from those presented in the main document (See Supplemental S2).

## Results

### Participant Inclusion

Owing to delays in receiving a medical cannabis card, two participants from the MCC group completed the majority (> 45 / 90 days) of their daily diary assessments prior to receiving the MCC card and were excluded from primary analyses. Additionally, to balance result generalizability, which may be undermined by strict thresholds for participant inclusion (Ji *et al*., 2018), and the complexity of missing data and model stability, we used a liberal criterion for completion rate of daily assessments for inclusion in our primary analyses. As a result, only three participants who did not have at least 9 out of 90 (10%) days with completed assessments were excluded from primary analyses. Therefore, our final primary analytic sample was 181 participants. Daily diary assessment completion rates were not significantly associated with primary study variables and are presented with (N=186) and without (N=181) excluded participants (see below).

### Daily Diary Completion Rates

Completion rates of daily surveys were high overall and consistent with prior ambulatory assessment studies of cannabis use in clinical samples with shorter study periods (e.g., less than 30 days (Goodhines *et al*., 2019)(Verdoux *et al*., 2003)(Buckner *et al*., 2012) see discussion). Among the full sample (N=186; no analysis-specific exclusions, see above), the median number of days with completed surveys was 72 out of 90 days, with a mean of 66.21 (SD=20.02) and range of 1-90 (Figure 1A). In the final analytic sample (N=181), the median number of days with completed surveys was 72 out of 90 days, with a mean of 67.00 (SD=18.68) and range of 12-90 (Figure 1A). Completion rates remained high throughout the entirety of the 90-day study period (see Supplemental S3).

**Figure 1.**
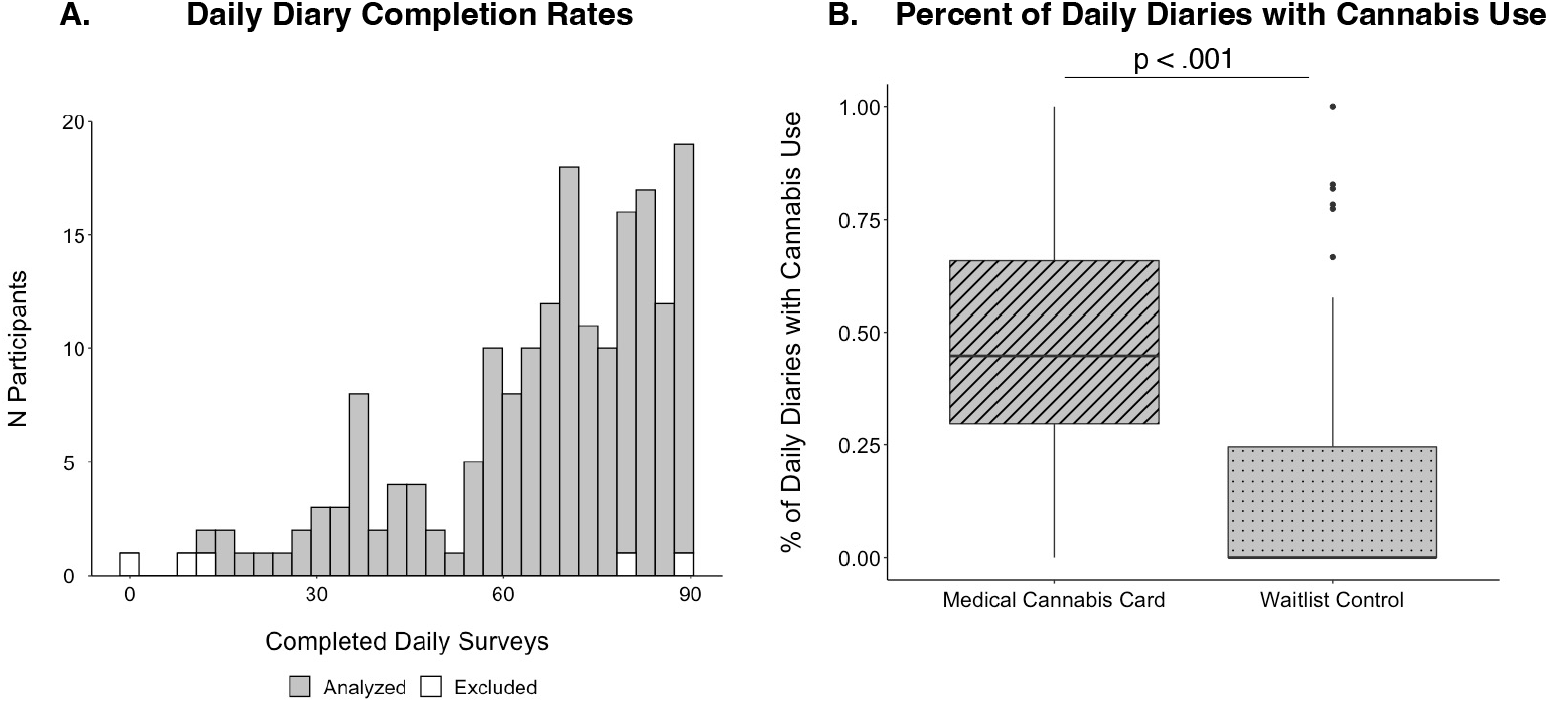
Daily Cannabis Diary Completion. A) Histogram displaying the number of completed diaries for each participant. White bars indicate the five excluded participants; grey bars indicate the 181 participants in the final analytic sample (see Methods). B) Boxplots displaying the percent of completed daily surveys where participants indicated they had used cannabis in the medical cannabis card (MCC) group compared to the waitlist control (WLC) group.

Among the full sample (N=183), the total number of completed surveys did not significantly differ between MCC (mean=68.45 days) and WLC (mean=63.31 days) groups (t=1.68, p=.095), nor as a function of presenting problem used for stratification in randomization (self-reported challenges with sleep, pain or mood; F=0.96, p =.386), and was likewise not related to baseline sleep (r= .005, p =.943), pain (r = .140, p =.186), depression (r = -.046, p =.529), or anxiety (r =-.103, p =.160) measures. The number of completed surveys did not significantly differ between men and women (t=0.98, p=.327). There were significant associations between the number of completed surveys and participant age (r = .237, p =.001), where older participants completed more surveys, and completed surveys and years of education, where those with more years of education completed more surveys (r = .175, p =.017). Age and years of education were used as covariates in all subsequent analyses. The pattern of significant associations between completion rates of daily surveys and study variables was unchanged when restricting analyses to only the participants in the primary analytic sample (N=181; see Methods for inclusion criteria).

### Medical Cannabis Card Group Reports More Cannabis Use Days than Waitlist Control

Daily diary data confirm randomization to a medical cannabis card (MCC) is associated with more frequent subsequent cannabis use; the MCC group had a significantly higher percentage of daily reports that included cannabis use, compared to the WLC (MCC: 48.2% vs WLC: 15.4%, p < .001)(Figure 1B). Daily diary data likewise indicated a significant increase in cannabis frequency as a function of time since the baseline visit in the MCC group (p=.007), but not the WLC group (p=.071) (see below).

### Validating Daily Diary with Field-Standard Interview Assessments and Urinalysis

To validate cannabis use data collected via web-based, daily diaries, we compared the MCC group’s (n=101 [1 MCC participant did not have 1 month follow-up data]) cannabis use frequency from this method (percentage of daily surveys reporting cannabis use) with 1) cannabis frequency identified in a field-standard, in person interview-based assessment (see Methods) querying the first month of the daily diary period and 2) the presence of cannabinoid metabolites in urine after the first month of the daily diary period (i.e., urinalysis from experimenter-administered, two-week or one-month visits). The first month of data were used in these analyses given the potential lingering effects of cannabinoid metabolites in urine following use, and to avoid “carry-over” effects where qualitatively positive urinalysis results from the first month might influence subsequent results (cf., (Schuster *et al*., 2020). Qualitative results (detected versus non-detected) were used given the high degree of individual variability in metabolite detection from urinalysis that is dependent on person-specific and methodological factors ((Goodwin *et al*., 2008)(Gilman *et al*., 2021b) see Supplemental S1 for additional discussion).

Cannabis use frequency collected via daily diaries (percentage of daily surveys with cannabis use) over the first month of the study was robustly associated with retrospective past month cannabis use frequency (number of days used) determined via structured interview questions with study staff at the experimenter-administered, four-week visit (standardized regression coefficient: β = .685, p < .001, partial R^2^=.468 while covarying participant age and years of education) (Figure 2A). Cannabis use frequency collected via daily diaries also disambiguated those in the MCC group with any cannabinoid metabolites (see Supplemental S1 for complete metabolite list) in their urine compared to those without detectable cannabinoids in their urine at either the two week or one month experimenter administered (cannabinoid metabolite detected after first month [n=75]: median diaries with use=46.3 %, cannabinoid metabolites not detected after first month [n=26]: median diaries with use = 13.0%, p < .001)(Figure 2B) as well as specifically those with THC metabolites (THC metabolite detected after first month [n=70]: 46.7 % diaries with use, THC metabolites not detected after first month [n=31]: 21.4% diaries with use, p < .001). See Supplemental S1 and (Gilman *et al*., 2021b) for extended detail on urinalysis scoring procedures.

**Figure 2.**
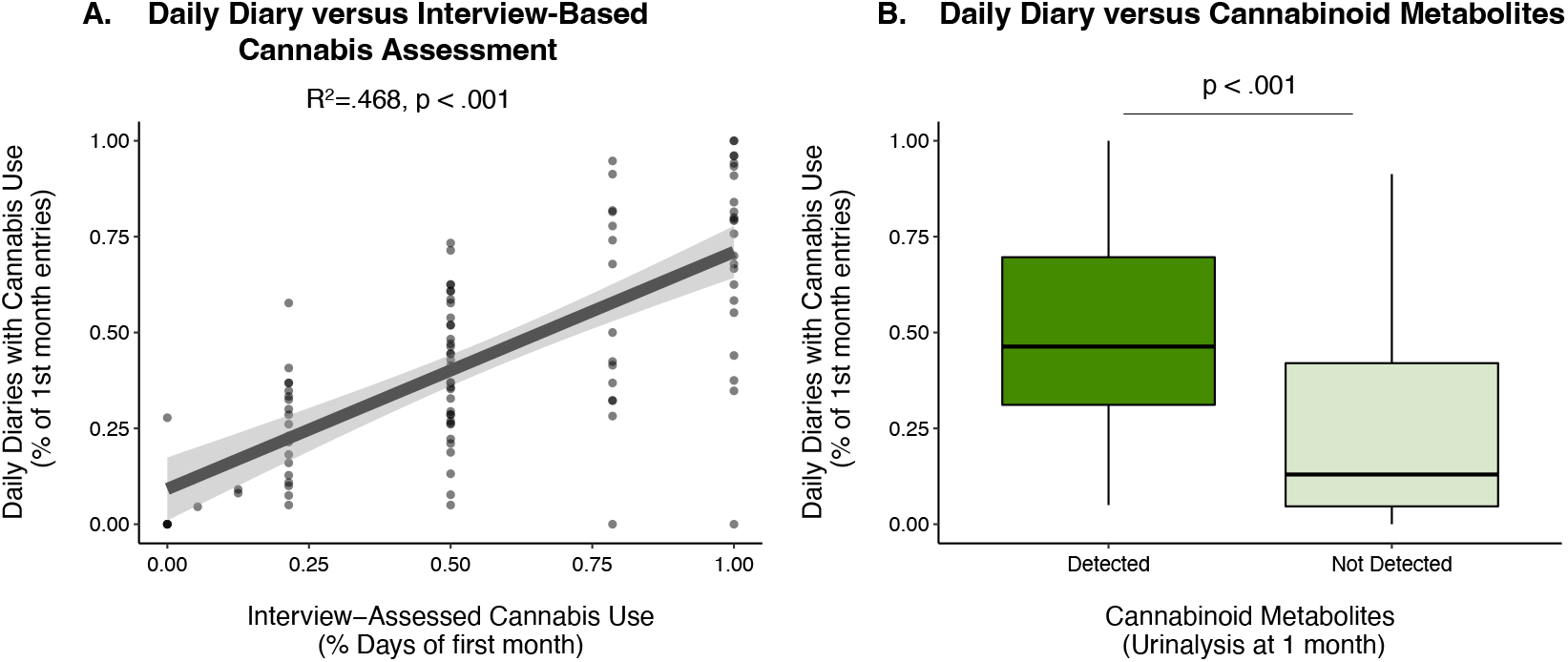
Daily Cannabis Diary Validation. A) Association between per-participant past month cannabis use frequency assessed via daily diary (y-axis) and via experimenter interview (x-axis) among those participants in the medical cannabis card (MCC, n=101) group. B) Per-participant past month cannabis use frequency derived via daily diary for those participants in the MCC with and without cannabinoid metabolites detected in their urine.

### Same Day Associations between Medical Cannabis Use and Health Symptoms in the MCC Group

Having established the feasibility and validity of the web-based daily cannabis assessments, we next examined within-person associations between cannabis use and health symptoms across the full 90-day monitoring period. Among the full MCC group (N=102), better sleep quality was reported for the night following cannabis use days compared to nonuse days (difference in standard deviation units: Δz = .115, p < .001)(Figure 3A). As our primary analyses utilized within-person day-level cannabis use and health symptom information and covaried for between-person associations between cannabis frequency and sleep (as well as salient demographic variables: see Methods), this result is consistent with medical cannabis use being associated with small, but significant same day improvements in self-reported sleep quality. The effect of same-day cannabis use on sleep quality significantly varied (omnibus test statistic for interaction from mixed model: χ2 = 7.32, p = .026) as a function of participants’ presenting problem at baseline (self-reported challenges with sleep, pain or mood), with post-hoc testing demonstrating significant, same day improvements of sleep on cannabis use days, compared to nonuse days for participants who entered the study based on self-reported problems with sleep (Δz = .178, p = .007) and mood (Δz = .178, p < .001) but not pain (Δz = .022, p = .623)(Figure 3A). The effect sizes and statistical significance of associations between cannabis use days vs. nonuse days with sleep quality did not change when restricting the MCC sample to only those with cannabinoid metabolites detected via urinalysis (Supplemental S2). Cannabis use days and nonuse days did not statistically differ in self-reported pain (Δz = .023, p = .428)(Figure 3B) and interactions between presenting problem (self-reported challenges sleep, pain, or mood) and cannabis use vs. nonuse days were not significant with respect to pain symptoms (χ2 = 2.44, p = .295). There was a very small, but statistically significant difference between cannabis use days and nonuse days in the full MCC sample (Δz =-058, p = .026), suggestive of use days being associated with slightly lower depressive symptoms compared to nonuse days; Figure 3C). This effect however was not significant in supplemental analyses (S2) that restricted the sample to those with cannabinoid metabolites detected via urinalysis (Δz =-.038, p = .197). The interaction between presenting problem and use vs. nonuse days was not significant with respect to depressive symptoms in the full MCC sample (χ2 = 4.32, p = .115) or in those with cannabinoid metabolites detected via urinalysis (χ2 = 2.37, p = .306).

**Figure 3.**
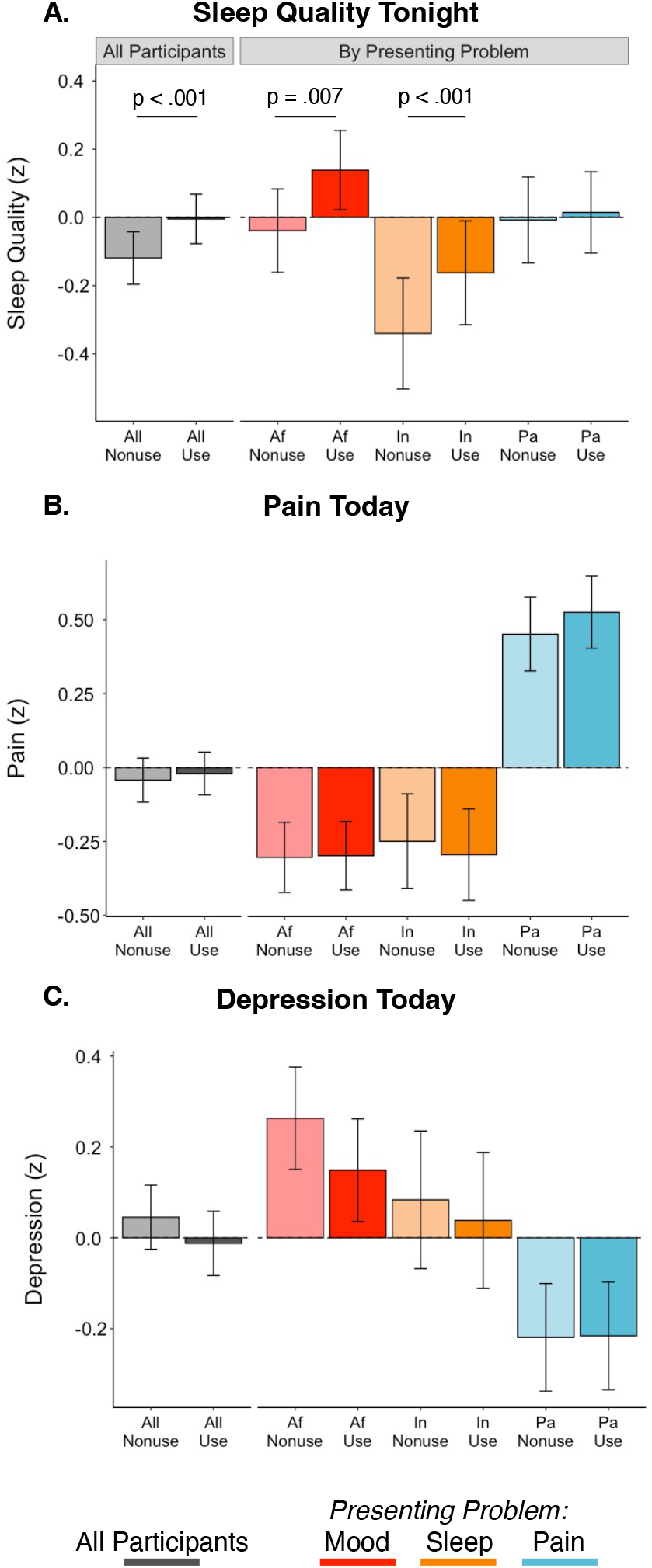
Differences in Health Symptoms between Cannabis Use Days and Nonuse Days in the Medical Cannabis Card Group. Differences in self-reported same night sleep quality (A), same day pain (B), and same day depression symptoms (C) in cannabis use days (darker colors) and nonuse days (lighter colors) in the medical cannabis card (MCC) group. Values shown for all MCC participants (N=102; grey) and those whose presenting problem was mood (depression or anxiety symptoms, Af: n=44, red), challenges with sleep (In: n=22, orange), or pain (Pa: n=36, blue). Displayed values are estimated marginal means and their standard errors from linear mixed effects models fit in primary analyses (covariates include per-participant estimates of age, years of education, and the proportion of daily diary days that included cannabis use [i.e., between-person effect]); see Methods for more detail on parameterization).

### Linking Daily and Long-Term Sleep Self-Reported Sleep Quality Changes Associated with New Medical Cannabis Use

Having demonstrated that compared to nonuse days, cannabis use days were associated with small to moderate statistically significant improvements in self-reported sleep quality including in analyses leveraging repeated urinalysis to confirm cannabis reports, we next sought to determine how these same day sleep improvements may manifest in long-term changes in sleep across the duration of the daily diary period. We found that across the daily diary period (90 days), the MCC group displayed significant aggregate increases in self-reported sleep quality following randomization (per-day standardized association (z units): β = .002, p =.007; total sleep change across 90 days (z units): .195), while the WLC group did not (per-day standardized association (z units): β = -.0003, p=.652; total sleep change across 90 days (z units): .030) (Figure 4A). This effect was driven by those in the MCC group with a presenting problem of insomnia, whose sleep quality significantly increased over time (per-day standardized association (z units): β = .006, p<.001; total sleep change across 90 days (z units): .675) and was significantly different (χ2 = 11.45, p =.003) than MCC participants with a presenting problem of pain or mood (Figure 4A). The pattern of longer-term increases in sleep quality across the daily diary period among those with a presenting problem of insomnia was mirrored in being the only MCC subgroup with a significant increase (per-day odds ratio: 1.02, p<.001; probability of a cannabis use day on study day 1: .370, probability of use day on study day 90: .832) in the frequency of cannabis use days following randomization (Figure 4B). This suggested that long-term improvements in sleep-quality were likely driven by an increase in cannabis use frequency, rather than for example, the lasting effect of a single cannabis use.

**Figure 4.**
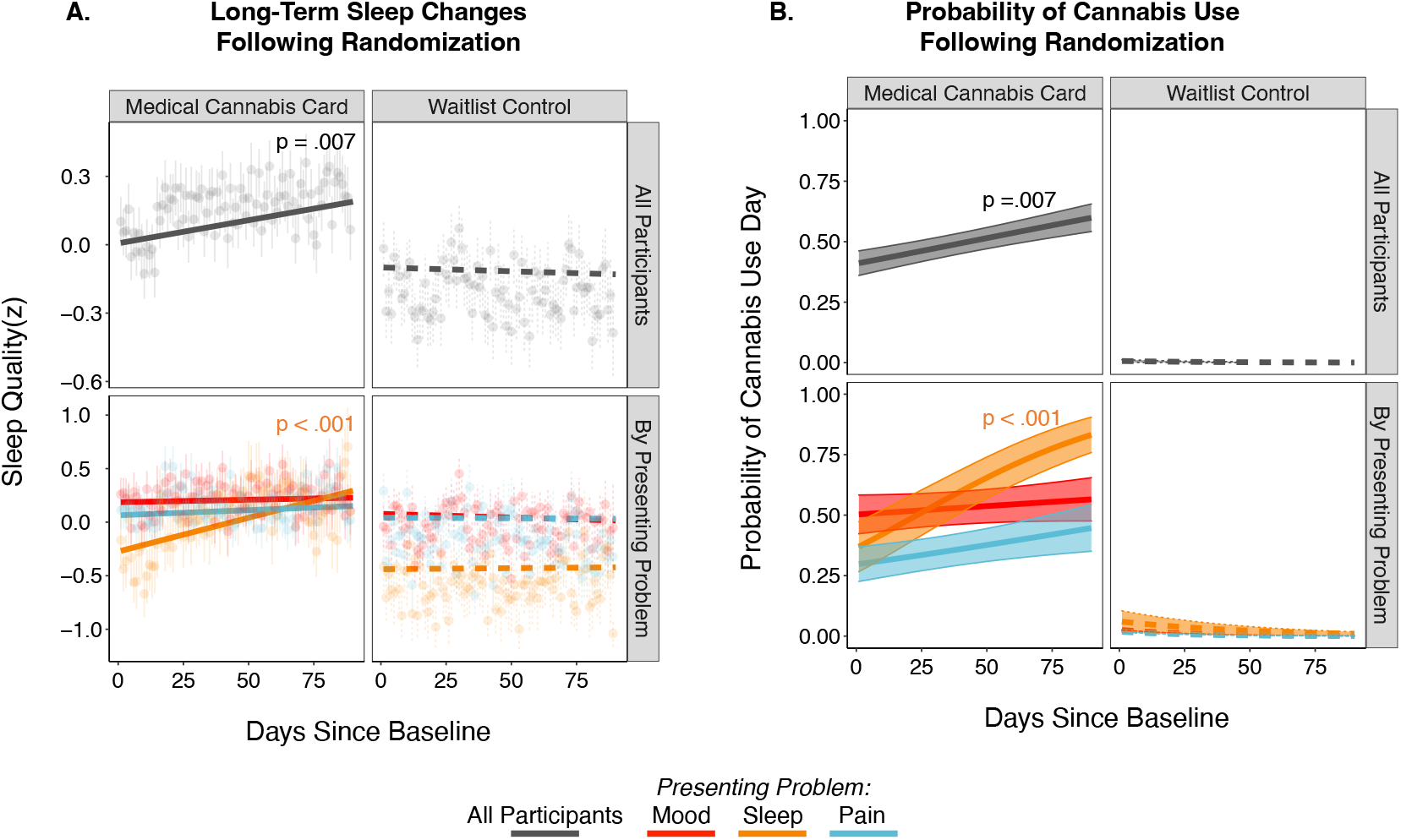
Duration of Cannabis’ Effect on Self-Reported Sleep. A) Sleep quality changes across the 90-day daily diary period for medical cannabis card (MCC, n=102; left) and waitlist control (WLC, n=79; right) groups shown for all participants in each group (top row) and separately by presenting problem (bottom row). Note, fits and data are averaged across use days and nonuse days. Data points are cross-participant means and standard errors of raw data; model fit lines are adjusted for random effects and primary model covariates (per-participant age, years of education, and proportion of daily diary days that included cannabis use; see Methods for more detail on parameterization). B) Probability of a cannabis use day across the 90-day daily diary period for the MCC (left) and WLC (right) groups for all participants (top row) and by presenting problem (bottom row).

## Discussion

This project establishes the feasibility and validity of a daily diary, experience sampling design for assessing medical cannabis use. With this method, the project demonstrates that cannabis use is associated with same day improvements in self-reported sleep quality, but not depressive, or pain symptoms. Through the parent project’s structure as a pragmatic randomized trial of medical cannabis cards and concurrent use of objective, urinalysis measures of cannabis use, this work also provides convergent evidence to mitigate key potential confounding inter-individual difference variables (e.g., variables equivalent across randomization groups: age, education, sex) and reporting bias, respectively, that may have otherwise accounted for the observed associations between same day cannabis use and sleep quality. Nevertheless, the improvement of sleep quality in those assigned to MCC occurred in the context of increasing frequency of cannabis use, suggesting those using cannabis to address problems with sleep should use caution, as more frequent cannabis use is not without risk and could lead to cannabis use disorder (Gilman et al., invited to resubmit, (Gilman *et al*., 2021a)).

### Feasibility and Validity of Daily Diary Design in Medical Cannabis Studies

Completion rates of daily surveys were high across our long-term (90 days) daily diary design (80% of surveys were completed on average) and approximately equivalent to other, far shorter experience sampling studies of cannabis use (e.g., >1 month 60-90% completion rates (Goodhines *et al*., 2019)(Verdoux *et al*., 2003)(Buckner *et al*., 2012). Using per-participant and group-level analysis procedures, we also found that completion rates remained within this range throughout the study period (Supplemental S3). We did find small, but significant associations suggesting older participants and those with higher education had higher daily survey completion rates. Given the novelty of the current study design for those seeking medical cannabis however, future work should seek replication of these associations. The small magnitude of the associations between completion rates and demographic factors and high overall completion rate in this study nevertheless suggest broad feasibility of this daily diary design of medical cannabis uses.

The current work also validates the daily diary, experience sampling method for medical cannabis, as cannabis use frequency derived from daily surveys well-aligned with field-standard, interview-based cannabis assessment as well as cannabinoid metabolite detection from urinalysis. To our knowledge, this is the first validation of daily, web-based cannabis use metrics with field standard interview assessments and urinalysis in medical cannabis use. Moreover, even among the rapidly increasing number of experience sampling studies of recreational cannabis use, few studies have sought validation of web-based cannabis assessments. While existing evidence has already suggested general validity of self-reported cannabis frequency (Martin *et al*., 1988) (although questions remain for cannabis dose and potency (van der Pol *et al*., 2013)), the current validation of daily web-based cannabis assessments provides essential clarification and support of modern experience sampling studies of cannabis use.

Feasible and valid web-based assessments of medical cannabis use are well-suited to current, highly variable real-world patterns of medical cannabis use that lack federal regulation, physician oversight, and clear guidelines on dosing. Such daily diary assessments of medical cannabis may thus capture both real-world patterns of cannabis use in future pragmatic clinical trials and also compliment formal efficacy trials to monitor study adherence and collect additional day-to-day or moment-to-moment use patterns. Web-based daily diary cannabis assessments will also be useful in future research that integrates photography or video to better remotely document cannabis product labels and ultimately estimate dosage, in efforts to address current lack of standards in dosage estimates.

### Medical Cannabis is Associated with Same Day Improvements in Sleep Symptoms

Using the validated daily diary design, we demonstrate that medical cannabis use is associated with an improvement in same night sleep quality. The observed improvement in sleep quality on cannabis use days, compared to nonuse days, was small-to-moderate (Δz ∼.18) with respect to current effect size benchmarks (typical psychology effect sizes ranging between r=.11 and r=.29 (Gignac and Szodorai, 2016)) and is considered to be practically (Funder and Ozer, 2019) and likely clinically (Rutledge and Loh, 2004) meaningful. This is notable as such small-to-moderate improvement is observed on the level of single days, instead of inter-individual differences or long-term change over the course of many weeks in which effect sizes are often interpreted (see (Gabriel *et al*., 2019)for more discussion). Same day sleep improvements among new medical cannabis users is consistent with prior work in a community sample of college students during a shorter monitoring period (14 days) that demonstrated self-reported same day sleep improvements among recreational cannabis users (Goodhines *et al*., 2019). These same day, within-person effects of cannabis-related sleep improvements are further consistent with a meta-analysis of sleep outcomes from clinical trials of therapeutic cannabinoids (Abrams, 2018) that found an overall small effect size, with 11 out of 19 trials reporting improvements in sleep quality or sleep disturbances

Supporting clinical research, basic science highlights an essential role of the endogenous cannabinoid system in sleep (see (Prospéro-García *et al*., 2016)(Babson *et al*., 2017) for review). Consistent with a potential direct role of cannabis use on sleep, this prior research has particularly implicated the CB1 receptor (Mechoulam *et al*., 1997)(Prospéro-García *et al*., 2016), where the primary psychoactive component of cannabis, Δ9-tetrahydrocannabinol (THC) is a partial agonist (Pertwee, 2008). In the context of emerging indirect, observational, and open-label/single-blinded clinical research and established basic science, expanded research investigating cannabinoids as a treatment for sleep challenges is warranted. The success of the current project utilizing an experience sampling, daily diary design suggests such treatment research may optimally utilize flexible, ecologically valid designs together with expanded and more objective sleep assessments within these ecologically valid designs (e.g. actigraphy watches).

### Medical Cannabis is not Associated with Same Day Improvements in Pain or Mood Symptoms

We did not find same day improvements in pain or depressive symptoms, which is consistent with our prior work looking at 12-week changes in health symptoms from in-person monthly assessments (i.e., non-daily diary) in adults randomized to receive a medical cannabis card (e.g. Gilman et al., invited to resubmit, (Gilman *et al*., 2021a)). Chronic pain is one of the most common complaints leading to an interest in medical cannabis use (Reinarman *et al*., 2011) and while some preclinical models suggest cannabinoids may regulate pain (Woodhams *et al*., 2015), clinical results have been inconclusive (Haroutounian *et al*., 2021)(National Academies of Sciences and Medicine, 2017)(Mücke *et al*., 2018). The current project finds no substantial evidence of cannabis improving same day pain symptoms. Given the success of the daily diary method for medical cannabis in the current project and established intraindividual variability in chronic pain symptoms (Mun *et al*., 2019) (O’Brien *et al*., 2011), however, if pursued, future work may utilize experience sampling designs to further explore same day associations in refined samples with expanded metrics.

Depressive symptoms are also associated with increased cannabis use in epidemiological samples (Onaemo *et al*., 2020)(Degenhardt *et al*., 2003) and are frequently cited as a reason to pursue medical cannabis (Reinarman *et al*., 2011). The current project however did not find substantive improvement in self-reported depressive symptoms among adults randomly assigned to access to medical cannabis use, which is largely consistent with existing evidence (Abrams, 2018). While there was one very small, but statistically significant difference in use versus nonuse days, this relationship was no longer observed when restricting the sample to those with verified cannabinoid metabolites. The high interest in use of cannabis for depressive symptoms together with the lack of improvement in depression symptoms among those using medical cannabis is concerning given that those with affective disorders (e.g., depression, anxiety disorders) are likely at significantly increased risk for developing a cannabis use disorder (Onaemo *et al*., 2020) Additional work suggests heavy cannabis use may increase risk for depression (Smolkina *et al*., 2017)(Lev-Ran *et al*., 2014) and other psychiatric illnesses (Livne *et al*., 2022)(although also see (Haney and Evins, 2016)), particularly among adolescents and young adults who often use cannabis in the context of normative, functional brain changes (Luna *et al*., 2015; Tervo-Clemmens *et al*., 2020). Taken together, it will be important for future medical cannabis treatment research to carefully assess symptoms of cannabis use disorder during treatment course and evaluate for exacerbation in depressive and other psychiatric symptoms.

### Limitations

This project is highlighted by several strengths, including randomization of participants to medical cannabis card access, a novel daily diary, experience sampling design, biochemical and interview-based validation of daily cannabis surveys, and a relatively large sample size for the number of repeated assessments (e.g., 181 participants with a median number of 72 assessments per-participant). There are, however, limitations worth noting. First, the current project relied on exploratory analyses of daily self-reported single-item assessments of sleep, pain, and mood symptoms that were included in the design to minimize participant burden and maximize completion over the very long study period (90 daily diary days). It is however essential for future work to consider replicating these results with objective measures and validated, clinician-blinded ratings. Another potential limitation of the current project is the likelihood of a high degree of variability of cannabis route of administration, dose, and potency among participants using medical cannabis. In part, the current, pragmatic study of medical cannabis was designed to test for effectiveness given these variable, real-world conditions, where participants are permitted to make their own decisions regarding cannabis products and dosing.

While we performed sensitivity analyses to ensure that the results were unchanged when restricting analyses to those with cannabinoid metabolites detected in their urine, lack of daily information on dosing, route of administration, and potency, prevent clear conclusions regarding pharmacological effects of cannabis. The success of the long-term web-based daily cannabis surveys and experience sampling design however, suggests future work with expanded objective measures of cannabis dose and potency (e.g., through regular laboratory-based testing) may be integrated with ecologically valid data collection on day-to-day use patterns and health symptoms.

### Conclusion

This project establishes the feasibility and validity of integrating a daily diary design into a pragmatic clinical trial of medical cannabis. Within this design, exploratory analyses support same-day improvements in sleep, but not pain or mood symptoms. Future work with expanded and objective measures of cannabis use and health symptoms, including indices of cannabis use disorder, can build upon the ecologically valid longitudinal design.

## Data Availability

All data produced in the present study are available upon reasonable request to the authors.

## Data Sharing Statement

All data, code, and materials used in the analyses can be provided by Brenden Tervo-Clemmens, Jodi Gilman, and Massachusetts General Hospital pending scientific review and a completed data use agreement/material transfer agreement. Requests for all materials should be submitted to Brenden Tervo-Clemmens and Jodi Gilman.

---

**Supplemental S1**. Cannabinoid Quantification from Urinalysis.

The quantification and detection of cannabinoid metabolites from urinalysis for this sample has been previously presented (Gilman *et al*., 2021b). We used identical scoring procedures as detailed in this prior work. Owing to the high degree of individual variability in metabolite detection from urinalysis that is dependent on person-specific (e.g., route of administration, hydration, metabolism and excretion rates) and methodological (e.g., assay sensitivity, specificity, and accuracy) factors (Goodwin *et al*., 2008), we used the composite metrics from (Gilman *et al*., 2021b) that coded whether urine samples contained any cannabinoid metabolites or any THC metabolites. Samples were tested for the following THC metabolites: 11-hydroxy-Δ9-tetrahydrocannabinol, 1-nor-Δ9-tetrahydrocannabinol-9-carboxylic acid glucuronide, 11-Nor-9-carboxy-Δ9-tetrahydrocannabinol, 11-nor-9-carboxy-Δ9-tetrahydrocannabivarin, Δ9-tetrahydrocannabinol glucuronide, Δ9-tetrahydrocannabivarin, as well as the following additional cannabinoid metabolites: 6-alpha-hydroxy-cannabidiol; 6-beta-hydroxy-cannabidiol; 7-hydroxy-cannabidiol; (3R-trans)-cannabidiol-7-oic acid; cannabichromene; cannabidiol; cannabidiol glucuronide; cannabidivarin; cannabigerol; cannabinol.

**Supplemental S2.**
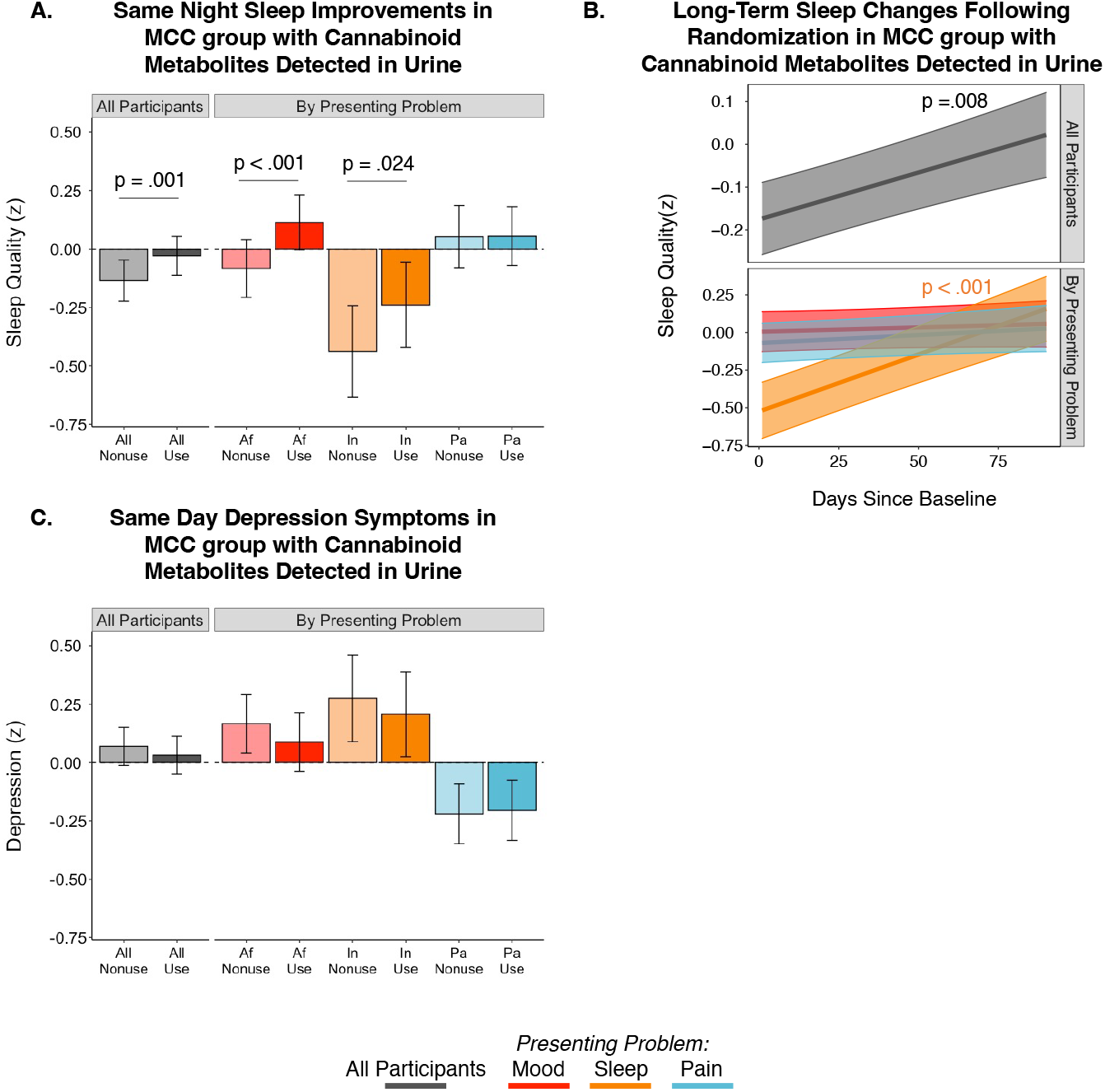
Sensitivity Analyses Restricting MCC group to those with Cannabinoid Metabolites Detected in their Urine During the Daily Diary Period. **A)** Differences in self-reported same night sleep quality (A in cannabis use days (darker colors) and nonuse days (lighter colors) in the medical cannabis card (MCC) group with cannabinoid metabolites detected in their urine (N=86; See main manuscript and S2 for more discussion). Values shown for all MCC participants (left) and those whose presenting problem was mood (depression or anxiety symptoms, Af: red), challenges with sleep (In: orange), or pain (Pa:blue). Displayed values are estimated marginal means and their standard errors from linear mixed effects models fit in primary analyses (covariates included the proportion of daily diary days that included cannabis use [i.e., between-person effect]; to improve model stability in this smaller sample and owing to the lack of predictive utility from participant age and years of education in primary models (p’s > .473), these covariates were removed from the models in these supplementary analysis. Inference regarding statistical significance was not changed with or without their inclusion. Models were otherwise identical to primary analysis: see Methods for more detail on parameterization. **B)** Sleep quality changes across the 90-day daily diary period for the MCC group with cannabinoid metabolites detected in their urine (N=86) shown for all participants (top row) and separately by presenting problem (bottom row). Note, fits are averaged across use days and nonuse days. **C)** Differences in self-reported same day depression symptoms in the medical cannabis card (MCC) group with cannabinoid metabolites detected in their urine (N=86) shown for all participants (left) and separately by presenting problem (right) as in (**A**).

**Supplemental S3.**
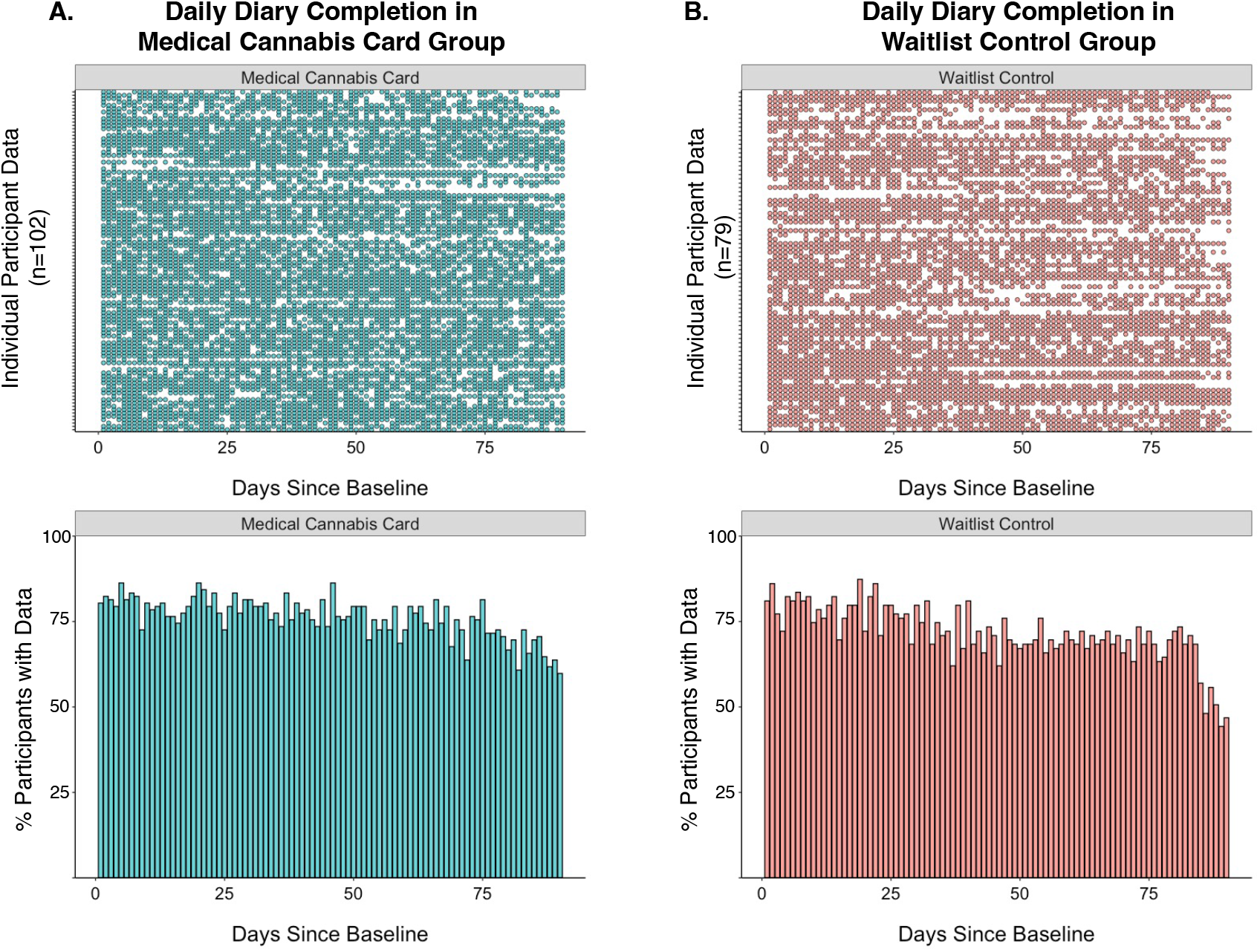
Visualization of Completed Daily Diaries Across the Study Period. **A)** Medical Cannabis Card group (n=102) daily diary completion. Top: visualization of per-participant daily diary completion, where each row corresponds to one participant across all 90 days of the study period; filled circles represent completed daily survey; open circles represent noncompleted/missing survey. Bottom: percent of participants with completed surveys for each of the 90 study days. **B)** Waitlist Control group (n=79) daily diary completion. Top: as in **A**, per-participant daily diary completion. Bottom: As in **A**, percent of participants with completed surveys for each of the 90 study days.

